# Undiagnosed COVID-19 in households with a child with mitochondrial disease

**DOI:** 10.1101/2022.03.21.22272358

**Authors:** Eliza M. Gordon-Lipkin, Christopher Marcum, Shannon Kruk, Elizabeth Thompson, Sophie E.M. Kelly, Heather Kalish, Kaitlyn Sadtler, Peter J. McGuire

**Affiliations:** Metabolism, Infection and Immunity Section, National Human Genome Research Institute, National Institutes of Health, Bethesda, MD; Data Science Policy, National Institute of Allergy and Infectious Diseases, National Institutes of Health, Bethesda, MD; Trans-NIH Shared Resource on Biomedical Engineering and Physical Science, National Institute of Biomedical Imaging and Bioengineering, National Institutes of Health, Bethesda, MD; Section on Immunoengineering, National Institute of Biomedical Imaging and Bioengineering, National Institutes of Health, Bethesda, MD

## Abstract

**Background:** The impact of the COVID-19 pandemic on medically fragile populations, who are at higher risk of severe illness and sequelae, has not been well characterized. Viral infection is a major cause of morbidity in children with mitochondrial disease (MtD), and the COVID-19 pandemic represents an opportunity to study this vulnerable population.

**Methods:** A convenience sampling cross-sectional serology study was conducted (October 2020 to June 2021) in households (N = 20) containing a child with MtD (N = 22). Samples (N = 83) were collected in the home using a microsampling apparatus and shipped to investigators. Antibodies against SARS-CoV-2 nucleocapsid (IgG), spike protein (IgG, IgM, IgA), and receptor binding domain (IgG, IgM, IgA) were determined by enzyme linked immunosorbent assay.

**Results:** While only 4.8% of participants were clinically diagnosed for SARS-CoV-2 infection, 75.9% of study participants were seropositive for SARS-CoV-2 antibodies. Most samples were IgM positive for spike or RBD (70%), indicating that infection was recent. This translated to all 20 families showing evidence of infection in at least one household member. For the children with MtD, 91% had antibodies against SARS-CoV-2 and had not experienced any adverse outcomes at the time of assessment. For children with recent infections (IgM^+^ only), serologic data suggest household members as a source.

**Conclusions:** COVID-19 was highly prevalent and undiagnosed in households with a child with MtD through the 2020-2021 winter wave of the pandemic. In this first major wave, children with MtD tolerated SARS-CoV-2 infection well, potentially due to household adherence to CDC recommendations for risk mitigation.

**Funding:** This study was funded by the Intramural Research Program of the National Institutes of Health (HG200381-03).

**Clinical trial number:** NCT04419870

## INTRODUCTION

Beginning in 2020, the COVID-19 pandemic, caused by the SARS-CoV-2 virus, has led to serious illness in hundreds of thousands of people in the United States and around the world.^1^ During this pandemic, individuals with preexisting medical conditions are at risk for adverse outcomes.^2^ The COVID-19 pandemic presents a unique opportunity to study the impact of a single viral infection on a medically vulnerable community. A prime example of a medically vulnerable community who has a deleterious relationship with infection is pediatric mitochondrial disease.

Deleterious variants in nDNA and mtDNA genes involved in mitochondrial function lead to disorders of oxidative phosphorylation, collectively known as mitochondrial diseases (MtD). MtD are the most common inborn errors of metabolism with a minimum prevalence of 1 in 5,000.^3^ The phenotype of MtD is multisystemic, involving organ systems with large energy requirements (e.g. central nervous system). Infection is a major cause of morbidity in children with MtD, precipitating acidosis and organ dysfunction with a rapidly fatal course.^4^ More than 80% of children with MtD may experience recurrent or severe infections^5^, and sepsis and pneumonia are common causes of death.^6^ Following respiratory viral infections, up to half of may experience life-threatening or neurodegenerative sequelae.^7^ As such, the COVID-19 pandemic poses a palpable threat to children with MtD, their caregivers and medical providers.

Public health messaging via the Centers for Disease Control (CDC) during the COVID-19 pandemic advised behaviors to mitigate transmission risk. Recently, we showed that families affected by MtD display high levels of adherence to these risk mitigation behaviors (RMBs) during the pandemic, likely due to fear of potential sequelae associated with infection.^8^ Despite these high levels of adherence amongst the MtD community, the home can still act as a hub for infection from household members^9^, many of which may be undiagnosed.^10^ As such, we hypothesized high penetrance of COVID-19 into households in the pediatric MtD community. To understand the status of pediatric MtD households and detect undiagnosed infections, we conducted a home-based serologic study. Our study allowed us to define the status of proximate contacts (i.e., family members) and children with MtD, as well as symptomatic and asymptomatic infection. The study was conducted through the 2020-2021 winter wave of the pandemic and is the first report of SARS-CoV-2 serologic status in this vulnerable population of patients.

## MATERIALS AND METHODS

### Patient cohort and clinical protocols

Participants self-identified their race and ethnicity from the following categories: American Indian or Alaskan Native, Asian, Black or African American, Hawaiian or Pacific Islander, Hispanic,

Non-Hispanic, White, more than 1, prefer not to answer, or other. The cohort of patients and their families (N=20 families) were self-selected participants in a natural history study of infection in children with MtD (ClinicalTrials.gov identifier: NCT04419870). Consent was obtained prior to study enrollment. This study was approved by the National Institutes of Health Institutional Review Board.

### Sample collection

At-home collection of capillary blood samples (30 μL) was performed using Mitra Specimen Collection Kit (Neoteryx, Torrence, CA) and a return shipping label. Upon receipt, microsamplers were stored dry at -80°C until elution and analysis. Corresponding clinical information was collected by telephone and encrypted online questionnaires.

### Serologic assays

The determination of SARS-CoV-2 antibodies from microsamples has been previously published.^10^ Briefly, whole blood samples were loaded onto 30 μL Neoteryx Mitra Microsampling device tips, dried, and stored in 500 μL Eppendorf tubes at -80°C. Microsampler tips were eluted in 500 μL of 1XPBS (Gibco) + 1% BSA + 0.5% Tween (Sigma-Aldrich, St. Louis, MO) at 4°C at 3,000 rpm. The eluate was stored at -80°C until use. Nucleocapsid, spike protein, and receptor binding domain antibodies were analyzed using enzyme linked immunosorbent assay.^10-12^ Longitudinal quality control and assay stability was ensured by the inclusion of controls for each plate (Recombinant anti-SARS-CoV-2 RBD, GenScript, and anti-SARS-CoV-2 Nucleocapsid, ThermoFisher Scientific, Waltham, MA). Seropositivity cut points were defined previously with thresholds based on the mean optical density (absorbance) plus 3 standard deviations.

### Statistics

Statistical analyses were performed using Microsoft Excel (Microsoft, Redmond, WA) and Graph Pad Prism (San Diego, CA). Data are presented as continuous variables, counts, percentages and means ± standard deviation. P < 0.05 was statistically significant.

## RESULTS

### Subject characteristics

Since the household is a risk factor for SARS-CoV-2 infection^9^, we collected capillary blood samples (N=83), from all resident members of the household (N=20 households) that contained a child with MtD. The study was conducted through the 2020-2021 winter wave of the pandemic. Two individuals from two separate families declined to participate, giving us 98% coverage of eligible participants. The characteristics of the children with MtD (N=22) are shown in Table 1. The mean age was 8.8 (±4.4 SD) years of age, with similar proportions of male (N=11) and female (N=11) subjects (Table 2). MtD diagnoses included: Leigh syndrome (N=11), Leigh-Like syndrome (N=4), mitochondrial depletion syndrome (N=1), MERFF (N=1), MELAS (N=2) and MtD not otherwise specified (NOS, N=3) (Table 1). Lesions of the central nervous system (a potential consequence of viral infection^7^), previously identified on MRI, were found in 82% of children with MtD. Participants represented 15 different counties across the United States, with 7-day case averages ranging from 1/100K population (Goodhue County, MN) to 73.9/100K population (Tarrant County, TX) at the time of sampling (Figure S1). Three families resided outside the United States where prevalence data were not available.

**Table 1:** Clinical characteristics of children with MtD. MtD = mitochondrial disease, LS = Leigh Syndrome, LLS = Leigh-like syndrome, mtDNA depl = mitochondrial depletion syndrome, MERRF = myoclonic epilepsy with ragged-red fibers, MELAS = mitochondrial encephalomyopathy, lactic acidosis, and stroke-like episodes, MtD NOS = mitochondrial disease not otherwise specified, CNS = central nervous system, BG = basal ganglia, BS = brain stem, CM = cerebellum, TH = thalamus, LD = leukodystrophy, MD with CNS = reported history of MRI findings, details unavailable.

| **MtD subject** | **Gene** | **Phenotype** | **CNS findings** |
| --- | --- | --- | --- |
| 1 | mt-ATP6 | LS | BG, BS, CM |
| 2 | SURF1 | LS | BS |
| 3 | NUBPL | LLS | BS, CM |
| 4 | mt-ND1 | LS | BG |
| 5 | mt-ND3 | LS | BG |
| 6 |  | mtDNA depl | NONE |
| 7 | mt-ATP6 | LS | BG, BS, CM, TH, LD |
| 8 | POLG | LLS | MD with CNS |
| 9 | TPK1 | LS | BG, TH |
| 10 | C12ORF65 | LS | BG, BS, TH, LD |
| 11 | mt-TL1 | MERRF | MD with CNS |
| 12 | AARS2 | MtD NOS | NONE |
| 13 | mt-TL1 | MELAS | NONE |
| 14 | mt-TL1 | MELAS | NONE |
| 15 | mt-ND5 | LS | MD with CNS |
| 16 | RMND1 | MtD NOS | LD |
| 17 | RMND1 | MtD NOS | LD |
| 18 | mt-CO1 | LLS | CM |
| 19 | EARS2 | LS | BS, TH |
| 20 | NDUFAF6 | LS | BG |
| 21 | IARS2 | LS | CM, LD |
| 22 | PDHA | LLS | BG, CM, LD |

**Table 2:** SARS-CoV-2 infection risk, diagnosis, and symptomatology. Caregivers of children with MtD (N=22) and household members ≥16 year of age (N = 49) were interviewed to determine risk factors and symptomatology for COVID-19. Data represented as counts with percentages or mean with standard deviation.

| Participants |  | Mt D | Household | P-value |
| --- | --- | --- | --- | --- |
|  |  | Number/total (%/Std dev) |  |  |
| Mean age |  | 8.8 (4.4) | 38.8 (11.2) |  |
| Sex |  |  |  |  |
|  | Males | 11/22 (50%) | 24/49 (49%) | 1.00 |
|  | Females | 11/22 (50%) | 25/49 (51%) | 1.00 |
| Job/school setting |  |  |  |  |
|  | On site | 2/22 (9.1%) | 8/49 (16.3%) | 0.71 |
|  | Hybrid | 2/22 (9.1%) | 9/49 (18.4%) | 0.48 |
|  | Remote | 18/22 (81.8%) | 23/49 (46.9%) | 0.01 |
|  | Not applicable | 0/22 (0.0%) | 9/49 (18.4%) |  |
| COVID19 exposure/testing/diagnosis |  |  |  |  |
|  | Exposed | 7/22 (31.8%) | 15/49 (30.6%) | 1.00 |
|  | Tested | 14/22 (63.6%) | 23/49 (46.9%) | 0.21 |
|  | Diagnosed | 1/22 (4.2%) | 3/71 (4.2%) | 1.00 |
| COVID19 symptoms |  |  |  |  |
|  | Fever or chills | 6/22 (27.2%) | 6/49 (12.2%) | 0.17 |
|  | New or worsening cough | 2/22 (8.3%) | 10/49 (20.4%) | 0.31 |
|  | New or worsening shortness of breath | 1/22 (4.2%) | 5/49 (10.2%) | 0.66 |
|  | Pneumonia | 2/22 (8.3%) | 1/49 (2.0%) | 0.23 |
|  | Muscle or body aches | 0/22 (0.0%) | 10/49 (20.4%) | 0.03 |
|  | Vomiting or diarrhea | 0/22 (0.0%) | 5/49 (10.2%) | 0.31 |
|  | Loss of taste or smell | 0/22 (0.0%) | 1/49 (2.0%) | 1.00 |

### SARS-CoV-2 infection risk, testing, and symptomatology

We next assessed risk factors for SARS-CoV-2 exposure (Table 2). During the COVID-19 pandemic, many school districts and workplaces adopted remote learning/work modalities as a risk mitigation strategy. In our cohort, the majority of children with MtD (81.8%) engaged in remote learning while for household members, remote work/school was lower (46.9%, P < 0.01) with more instances of onsite/hybrid scenarios where individuals may be in close contact to facilitate COVID-19 spread. Concern for exposure may be reflected in testing for SARS-CoV-2: 63.6% of children with MtD and 46.9% of household members received testing. Reported COVID-19 exposure occurred in 31.8% of children with MtD and 30.6% of household members. Despite these risks, only 4.8% individuals reported that they were diagnosed with COVID-19 prior to sample collection; one was a child with MtD. As such, the majority of the cohort had not been diagnosed by a healthcare professional.

To further assess potential SARS-CoV-2 infection, we asked about common COVID-19 symptoms since March 2020 (Table 2). The most common symptom in children with MtD was fever (27.2%), although this did not significantly differ in frequency from other household members (12.2%, P=0.17). The most common symptom reported by household members that contrasted with children with MtD was muscle aches (20.4% versus 0.0%, P=0.03). Overall, 31.8% of children with MtD and 49.0% of household members reported at least one symptom consistent with COVID-19. Despite the occurrence of these symptoms, there were no reports of hospitalizations, ICU admissions or deaths in any of the households.

### Serology

Following extraction from our microsampling apparatus, we performed serologic studies for IgM, IgG and IgA antibodies against viral nucleocapsid, spike protein, and receptor binding domain (RBD). IgM antibodies indicated recent infection, while IgG and IgA antibodies indicated past infection. Unvaccinated individuals were considered seropositive if they had antibody(ies) against any of the viral proteins tested. Individuals who were found to have nucleocapsid antibodies were considered seropositive regardless of vaccination status, since nucleocapsid antibodies cannot arise from vaccination. Overall, we found that 75.9% of participants were seropositive for at least one of the SARS-CoV-2 antibodies (Figure 1a).

**Figure 1:**
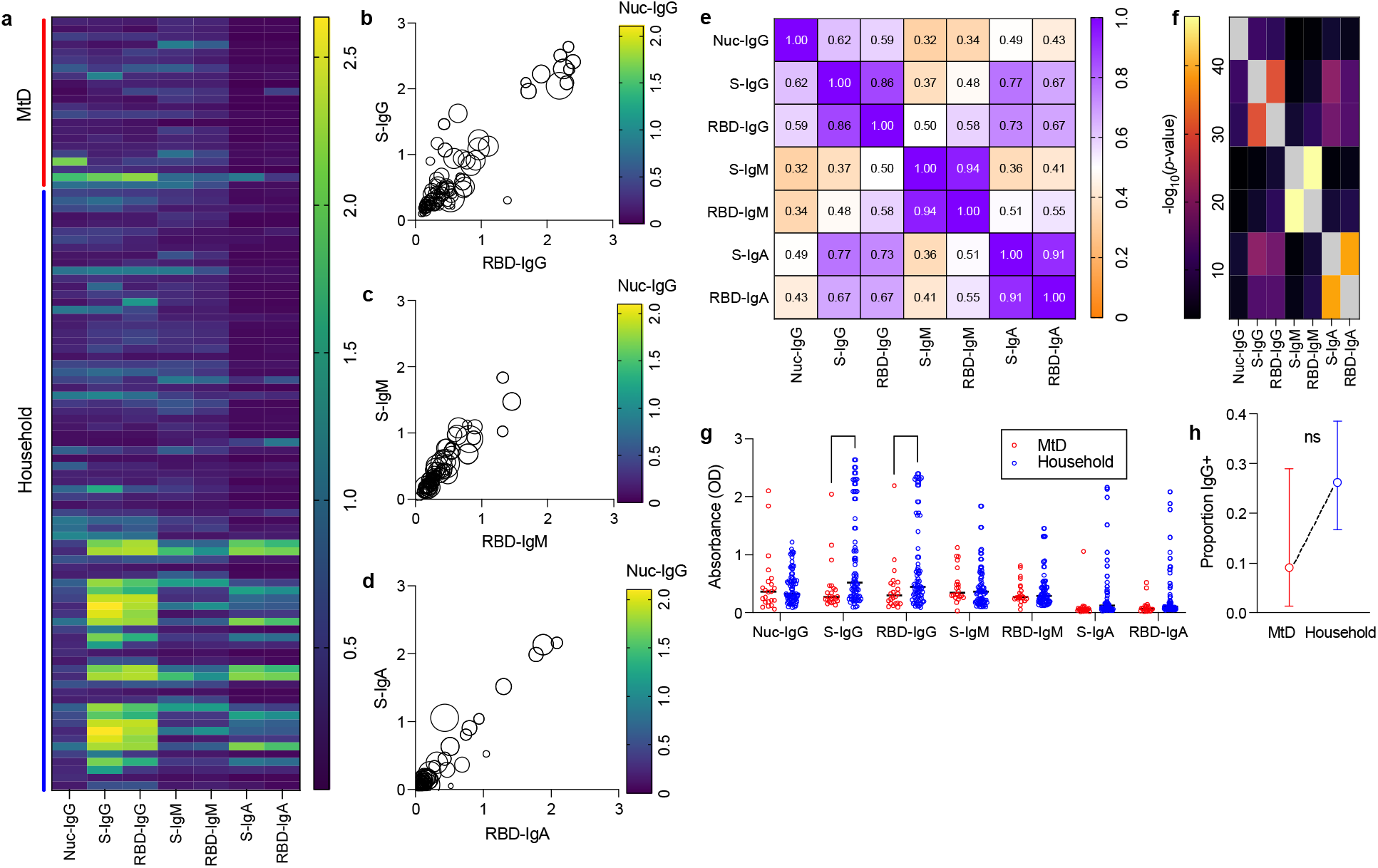
SARS-CoV-2 antibody profile in MtD patients versus family members. (a) Heatmap showing antibody prevalence in the blood of participants, Nuc = Nucleocapsid, S = Full Spike ectodomain, RBD = Spike Receptor Binding Domain. (b) Anti-spike IgG (c) Anti-spike IgM (d) Anti-spike IgA. (e) Spearman correlation matrix, two-tailed (f) P-values for spearman correlation. Y-axis labels are in Figure 1e. (g) Antibody analytes mean absorbance (OD) between MtD (red) and family (blue), significance = Two-way ANOVA with Tukey post-hoc correction for multiple comparisons *** = p <0.001, * = p < 0.05. (h) Proportion of participants that are IgG positive for spike or RBD protein.

We next looked for associations between the different antibodies tested. In general, absorbances for spike protein displayed a linear relationship with RBD (Figure 1b-d). Nucleocapsid IgG absorbance (circle color) lacked any association with spike and RBD absorbance. To further define relationships between antibodies, we constructed a correlation matrix (Figure 1e). There was a high degree of correlation between spike and RBD IgG (r = 0.86), IgM (r = 0.94) and IgA (r = 0.91), as expected, as the RBD constitutes a domain of the spike protein. Interestingly, absorbances for spike IgA also correlated with spike and RBD IgG (r = 0.77 and r = 0.73, respectively). All antibody absorbance correlations were found to be statistically significant (Figure 1f). We did not find any relationship between the presence of symptoms and the amount of antibody absorbance (data not shown). To capture potential differences in antibody responses, we compared the absorbances for each antibody between our two groups (Figure 1g). In general, children with MtD tended to have lower absorbances for spike and RBD IgG. When accounting for seropositivity, the proportion of individuals who were spike or RBD IgG^+^ tended to be lower for children with MtD, however, this finding was not statistically significant (Figure 1h).

The breakdown of antibody profiles for all unvaccinated individuals is shown in Table 3. For spike protein, the antibody profiles were similar for household members and children with MtD. A spike antibody profile of entirely spike IgM^+^ displayed a trend, being more frequent in children with MtD (20.9% versus 40.9%, P = 0.09). RBD antibody profiles were also similar for children with MtD and household members, except for IgM. An RBD antibody profile marked by entirely IgM^+^ was found more often in children with MtD (12.2% versus 36.4%, P = 0.03). These results suggest that subsets of children with MtD were experiencing more recent infections.

**Table 3:** Seropositivity profile in participants with anti-SARS-CoV-2 antibodies. IgG = immunoglobulin G, IgM = immunoglobulin M, IgA = immunoglobulin A, Spike = spike protein, RBD = receptor binding domain. Fisher’s exact test with P < 0.05 in bold.

|  | MtD | Household |  |
| --- | --- | --- | --- |
| Nucleocapsid | Number (%) |  | P value |
| *IgG*<sup>+</sup> | 2/22 (9.1%) | 5/49 (10.2%) | 1.00 |
| **Spike** |  |  |  |
| *IgG*<sup>+</sup> *IgM*<sup>+</sup> *IgA*<sup>+</sup> | 1/22 (4.5%) | 1/49 (2.0%) | 0.53 |
| *IgG*<sup>+</sup> *IgM*<sup>+</sup> | 0/22 (0.0%) | 2/49 (4.1%) | 1.00 |
| *IgG*<sup>+</sup> *IgA*<sup>+</sup> | 0/22 (0.0%) | 0/49 (0.0%) | 1.00 |
| *IgM*<sup>+</sup> *IgA*<sup>+</sup> | 0/22 (0.0%) | 1/49 (2.0%) | 1.00 |
| *IgG*<sup>+</sup> | 1/22 (4.5%) | 1/49 (2.0%) | 0.53 |
| *IgM*<sup>+</sup> | 9/22 (40.9%) | 10/49 (20.4%) | 0.09 |
| *IgA*<sup>+</sup> | 0/22 (0.0%) | 1/49 (2.0%) | 1.00 |
| Any *IgM*<sup>+</sup> | 9/22 (40.9%) | 14/49 (28.6%) | 0.41 |
| Any *IgG*<sup>+</sup> | 2/22 (9.1%) | 4/49 (8.2%) | 1.00 |
| **RBD** |  |  |  |
| *IgG*<sup>+</sup> *IgM*<sup>+</sup> *IgA*<sup>+</sup> | 0/22 (0.0%) | 0/49 (0.0%) | 1.00 |
| *IgG*<sup>+</sup> *IgM*<sup>+</sup> | 10/22 (45.4%) | 25/49 (51.0%) | 0.80 |
| *IgG*<sup>+</sup> *IgA*<sup>+</sup> | 0/22 (0.0%) | 0/49 (0.0%) | 1.00 |
| *IgM*<sup>+</sup> *IgA*<sup>+</sup> | 0/22 (0.0%) | 1/49 (2.0%) | 1.00 |
| *IgG*<sup>+</sup> | 2/22 (9.1%) | 7/49 (14.3%) | 0.71 |
| *IgM*<sup>+</sup> | 8/22 (36.4%) | 6/49 (12.2%) | **0.03** |
| *IgA*<sup>+</sup> | 0/22 (0.0%) | 0/49 (0.0%) | 1.00 |
| Any *IgM*<sup>+</sup> | 18/22 (81.8%) | 32/49 (65.0%) | 0.26 |
| Any *IgG*<sup>+</sup> | 12/22 (54.5%) | 32/49 (65.0%) | 0.43 |

To understand the risk posed to the child(ren) with MtD, we represented household structures as spoke diagrams with child(ren) with MtD at the center, parents at the top and in first position clockwise, followed by other family members (Figure 2). All 20 families had one or more individuals with evidence of infection. Two families, Family #13 and Family #15, had twins with MtD. For Family #15, both children were positive, while Family #13, had one positive child. Also of note is Family #17, where the affected child was the only positive household member. Eight children with MtD (Table 3 and Figure 2) from Families #1, #2, #4, #9, #11, #14, #15 (2 children), were found to be exclusively IgM^+^ for RBD, spike or both. Since IgM precedes the appearance of IgG, we next examined whether we could deduce potential transmission from other household members. In Families #1, #2, #4, and #9, both parents were IgG^+^. In Families #11 and #15, one of the parents was IgG^+^ for RBD, while another family member(s) was positive for IgG or IgA. Overall, all eight children with MtD had one or more household members who were IgG^+^, indicating that transmission may have originated from the household member.

**Figure 2:**
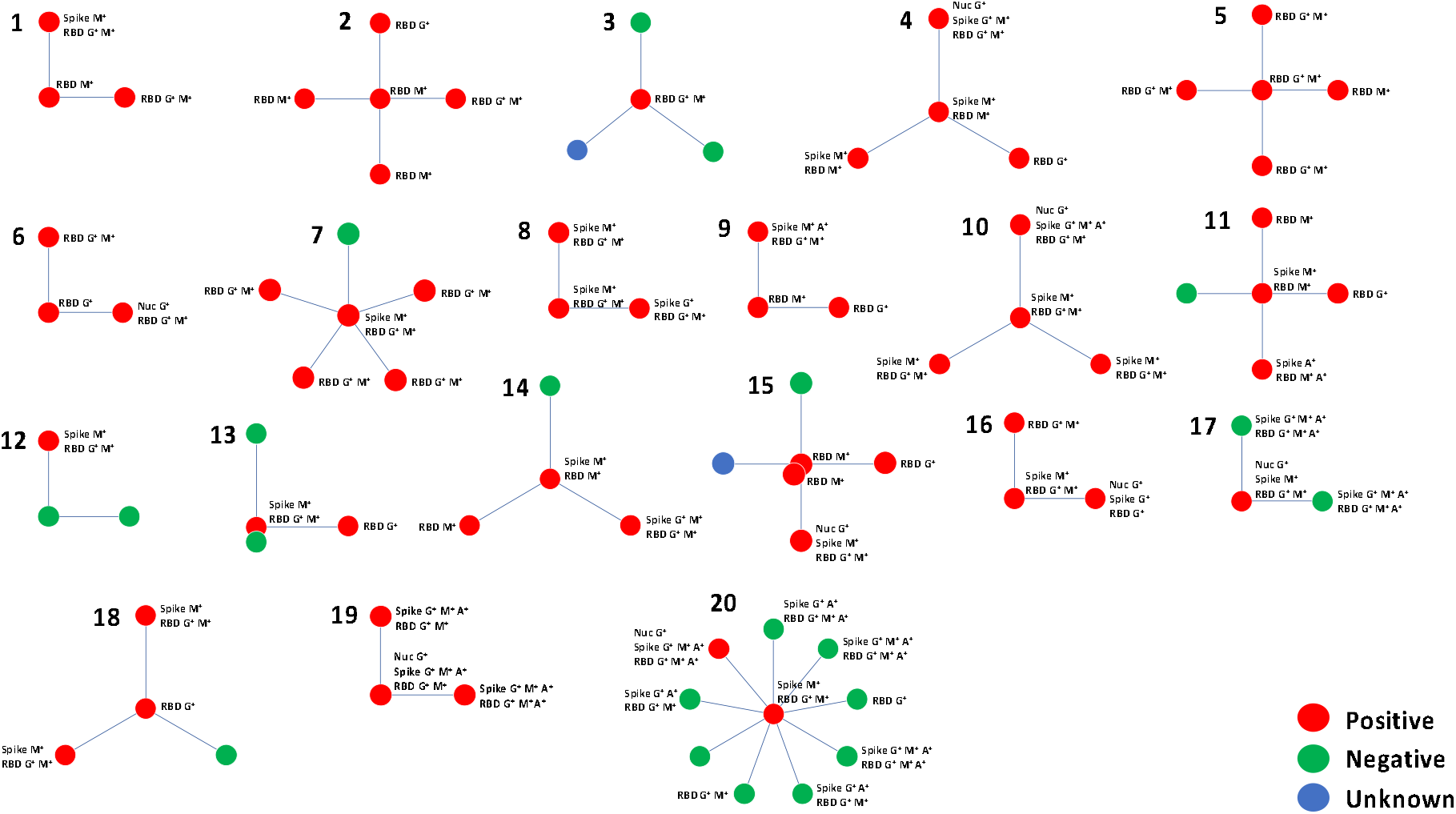
SARS-CoV-2 serology for families with a child with MtD. To highlight the relationship between individuals in the household and the child(ren) with mitochondrial disease are represented as a spoke diagram. The child(ren) with MtD is located at the center and is connected to household members by spokes. Viral nucleocapsid, spike protein, and receptor binding domain (RBD) serologies were performed as outlined in the methods. Samples were collected for each participant at a single timepoint surrounding the 2020-2021 winter wave of the COVID-19 pandemic. Families are numbered 1 through 20, and SARS-CoV-2 serology status is indicated by the color key. M^+^ = IgM antibodies, G^+^ = IgG antibodies, A^+^ = IgA antibodies, Spike = SARS-CoV-2 spike protein, RBD = receptor binding domain of the SARS-CoV-2 spike protein, Nuc = viral nucleocapsid.

Vaccinated members of the cohort (N=12) included Families #17, #19 and #20 (Figure 2). Antibody profiles for these individuals were consistent with vaccination. A few individuals, however, were still marked as seropositive for SARS-CoV-2 infection. Prior to vaccination, the parents in Family #19 were diagnosed with COVID-19 by their medical provider. A household member of Family #20, despite vaccination, was seropositive for IgG antibodies against the nucleocapsid, indicating past infection (Figure 2).

## DISCUSSION

The COVID-19 pandemic continues to present challenges to medically vulnerable pediatric communities. In addition to restricted access to medical care and services, there is also a persistent concern for adverse outcomes from SARS-CoV-2 infection. Children with chronic physical, developmental, behavioral or emotional conditions, disabilities, and those with medically complex conditions, are at increased risk for more severe illness and complications. Pediatric MtD is a complex medical condition with considerable morbidity associated with viral infection. Due to this potential deleterious relationship with infection, we considered this population a prime model for examining the impact of COVID-19 on a medically vulnerable pediatric community. Our results provide a previously undescribed view of the prevalence of the SARS-CoV-2 infection and outcomes in households of children with MtD; the majority of infections were undiagnosed and recent.

In general, our cohort displayed substantial penetration of SARS-CoV-2 into households. One explanation may be related to the prevalence of COVID-19 infection. At the height of the 2020-2021 winter surge, the 7-day average for new cases in the United States was >250,000.^13^ In addition to COVID-19 prevalence, household characteristics may have also played a role. Although virtual learning and telework became an important tool for helping maintain safe environments, not everyone was able to utilize these resources.^14^ In our study, over one third of household members ≥16 years of age reported onsite or hybrid work/learning scenarios, a risk factor for introducing viral infection. Indeed, employment-related exposure to SARS-CoV-2 can endanger not only workers, but also their household members.^15^

Another factor influencing the high degree of penetrance of SARS-CoV-2 may have been the limited availability of vaccinations. As vaccines are now becoming available for children, an added concern has emerged. While vaccination against SARS-CoV-2 can prevent infection, ameliorate COVID-19 severity^16^ and reduce household transmission^17^, there remains significant vaccine hesitancy for children with MtD due to the perceived risk of disease progression.^18^ The high degree of household seropositivity seen in our spoke diagrams combined with the vaccine hesitancy reported for children with MtD, suggests that pursuing household members for ring vaccination may be a worthwhile risk mitigation strategy. Ring vaccination is the targeting of individuals who are in contact with a population of concern for infection. In fact, ring vaccination has been considered as a strategy for the evaluation of vaccine efficacy and effectiveness.^19^ Future studies will not only need to address the uptake and efficacy/effectiveness of vaccination, but also the value of ring vaccination in the MtD community.

Similar to previous reports on the pediatric population ^20,21^, children with MtD did not experience any adverse outcomes during the study. Childhood factors that may account for this include: 1) Reduced ACE2 expression; 2) Frequent vaccinations sustaining immunity; 3) An abundance of natural antibodies, which foster a rapid immune response; 4) Cross-protection from infections with common cold coronaviruses. Another possibility is the high degree of adherence to RMBs in the MtD community, as mentioned above. This could result in low dose infection with oligo/asymptomatic disease and antibody production.^22^

Despite the absence of immediate serious outcomes, concerns still remain regarding the long-term consequences of host-pathogen interactions. Although unknown in MtD, evidence suggests that SARS-CoV-2 can impair mitochondria potentially leading to disease exacerbation.^23^ Individuals with COVID-19 can also exhibit neurologic signs, indicating CNS infection^24-26^; a finding supported by human iPSC cerebral organoid models and autopsy studies.^24,27^ Furthermore, CNS infection can persist, lasting up to 230 days.^24^ Overall, neuroinvasion of SARS-CoV-2 raises the concern for acute or chronic neurodegeneration via damaging neuroinflammation in children with MtD.^28^

Sources of potential bias are present in this study and should be taken into account when interpreting our results. All participants were identified via social media and family advocacy group advertising. This type of recruitment strategy preselects participants who have the resources to participate in social media. Inclusion criteria for study entry included molecular evidence of MtD via mtDNA depletion studies or the identification of a known pathologic variant. This may exclude individuals who do not have the health care resources to access genetic testing for disease diagnosis. Finally, the study was conducted through the 2020-2021 winter wave of the pandemic when the primary SARS-CoV-2 strain was the alpha variant. As the pandemic has progressed, newer variants have arisen (e.g. Delta and Omicron), which have different infection parameters and kinetics. While our study presents the only published SARS-CoV-2 serological study of children with MtD to our knowledge, larger and longer-term studies are needed.

In conclusion, our study highlights the number of undiagnosed cases and penetration of SARS-CoV-2 into a medically vulnerable community, children with MtD. More importantly, our understanding of the natural history of MtD, particularly the relationship with viral infection, has advanced. As the pandemic continues, future studies will help elucidate which viruses (or variants) are detrimental or well-tolerated, with the goal of improving clinical care. Generally speaking, our at-home sampling platform and study objectives serve as a template for overcoming research barriers and accessing understudied populations during and beyond the pandemic.

## Data Availability

All data produced in the present study are available upon reasonable request to the authors

## ACKNOWLEDGEMENTS

The authors would like to acknowledge the United Mitochondrial Disease Foundation and all of the families who participated in the study. We would like to acknowledge D. Esposito and FNLCR for providing the proteins used in serology. This work was supported by the Intramural Research Program of the National Institutes of Health including the National Human Genome Research Institute, the National Institute for Allergy and Infectious Diseases, and the National Institute for Biomedical Imaging and Bioengineering.

## Disclaimer

The NIH, its officers, and employees do not recommend or endorse any company, product, or service.

## REFERENCES

1. Team CC-R. Severe Outcomes Among Patients with Coronavirus Disease 2019 (COVID-19) - United States, February 12-March 16, 2020. MMWR Morb Mortal Wkly Rep 2020;69(12):343–346. DOI: 10.15585/mmwr.mm6912e2.

2. McMichael TM, Clark S, Pogosjans S, et al. COVID-19 in a Long-Term Care Facility - King County, Washington, February 27-March 9, 2020. MMWR Morb Mortal Wkly Rep 2020;69(12):339–342. DOI: 10.15585/mmwr.mm6912e1.

3. Ng YS, Turnbull DM. Mitochondrial disease: genetics and management. J Neurol 2016;263(1):179–91. DOI: 10.1007/s00415-015-7884-3.

4. Naviaux RK, Nyhan WL, Barshop BA, et al. Mitochondrial DNA polymerase gamma deficiency and mtDNA depletion in a child with Alpers’ syndrome. Ann Neurol 1999;45(1):54–8. (https://www.ncbi.nlm.nih.gov/pubmed/9894877).

5. Tarasenko TN, Pacheco SE, Koenig MK, et al. Cytochrome c Oxidase Activity Is a Metabolic Checkpoint that Regulates Cell Fate Decisions During T Cell Activation and Differentiation. Cell Metab 2017;25(6):1254–1268 e7. DOI: 10.1016/j.cmet.2017.05.007.

6. Eom S, Lee HN, Lee S, et al. Cause of Death in Children With Mitochondrial Diseases. Pediatr Neurol 2017;66:82–88. DOI: 10.1016/j.pediatrneurol.2016.10.006.

7. Edmonds JL, Kirse DJ, Kearns D, Deutsch R, Spruijt L, Naviaux RK. The otolaryngological manifestations of mitochondrial disease and the risk of neurodegeneration with infection. Arch Otolaryngol Head Neck Surg 2002;128(4):355–62. DOI: 10.1001/archotol.128.4.355.

8. Gordon-Lipkin E, Kruk S, Thompson E, et al. Risk mitigation behaviors to prevent infection in the mitochondrial disease community during the COVID19 pandemic. Mol Genet Metab Rep 2021:100837. DOI: 10.1016/j.ymgmr.2021.100837.

9. Madewell ZJ, Yang Y, Longini IM, Jr., Halloran ME, Dean NE. Factors Associated With Household Transmission of SARS-CoV-2: An Updated Systematic Review and Meta-analysis. JAMA Netw Open 2021;4(8):e2122240. DOI: 10.1001/jamanetworkopen.2021.22240.

10. Kalish H, Klumpp-Thomas C, Hunsberger S, et al. Undiagnosed SARS-CoV-2 seropositivity during the first 6 months of the COVID-19 pandemic in the United States. Sci Transl Med 2021;13(601). DOI: 10.1126/scitranslmed.abh3826.

11. Esposito D, Mehalko J, Drew M, et al. Optimizing high-yield production of SARS-CoV-2 soluble spike trimers for serology assays. Protein Expr Purif 2020;174:105686. DOI: 10.1016/j.pep.2020.105686.

12. Mehalko J, Drew M, Snead K, et al. Improved production of SARS-CoV-2 spike receptor-binding domain (RBD) for serology assays. Protein Expr Purif 2021;179:105802. DOI: 10.1016/j.pep.2020.105802.

13. Dong E, Du H, Gardner L. An interactive web-based dashboard to track COVID-19 in real time. Lancet Infect Dis 2020;20(5):533–534. DOI: 10.1016/S1473-3099(20)30120-1.

14. Vargo D, Zhu L, Benwell B, Yan Z. Digital technology use during COVID-19 pandemic: A rapid review. Hum Behav Emerg Tech 2021;3(1):13–24. (In English). DOI: 10.1002/hbe2.242.

15. Selden TM, Berdahl TA. Risk of Severe COVID-19 Among Workers and Their Household Members. Jama Intern Med 2021;181(1):120–122. (In English). DOI: 10.1001/jamainternmed.2020.6249.

16. Polack FP, Thomas SJ, Kitchin N, et al. Safety and Efficacy of the BNT162b2 mRNA Covid-19 Vaccine. N Engl J Med 2020;383(27):2603–2615. DOI: 10.1056/NEJMoa2034577.

17. Singanayagam A, Hakki S, Dunning J, et al. Community transmission and viral load kinetics of the SARS-CoV-2 delta (B.1.617.2) variant in vaccinated and unvaccinated individuals in the UK: a prospective, longitudinal, cohort study. Lancet Infect Dis 2021. DOI: 10.1016/S1473-3099(21)00648-4.

18. Gordon-Lipkin, Kruk S, Thompson E, et al. Vaccine Hesitancy toward the COVID19 Vaccine and Attitudes toward Ring Vaccination by Caregivers of Children with Mitochondrial Disease. Annals of Neurology 2021;90:S111–S111. (In English) (<Go to ISI>://WOS:000700182700137).

19. Kucharski AJ, Eggo RM, Watson CH, Camacho A, Funk S, Edmunds WJ. Effectiveness of Ring Vaccination as Control Strategy for Ebola Virus Disease. Emerg Infect Dis 2016;22(1):105–108. (In English). DOI: 10.3201/eid2201.151410.

20. Falahi S, Abdoli A, Kenarkoohi A. Claims and reasons about mild COVID-19 in children. New Microb New Infec 2021;41 (In English). DOI: ARTN 100864 10.1016/j.nmni.2021.100864.

21. Mallapaty S. Kids and COVID: why young immune systems are still on top. Nature 2021;597(7875):166–168. DOI: 10.1038/d41586-021-02423-8.

22. Hausdorff WP, Flores J. Low-dose and oral exposure to SARS-CoV-2 may help us understand and prevent severe COVID-19. Int J Infect Dis 2021;103:37–41. (In English). DOI: 10.1016/j.ijid.2020.11.171.

23. Ganji R, Reddy PH. Impact of COVID-19 on Mitochondrial-Based Immunity in Aging and Age-Related Diseases. Front Aging Neurosci 2020;12:614650. DOI: 10.3389/fnagi.2020.614650.

24. Chertow D, Stein S, Ramelli S, et al. SARS-CoV-2 infection and persistence throughout the human body and brain. Nature In Review 2021. DOI: 10.21203/rs.3.rs-1139035/v1.

25. Yates D. A CNS gateway for SARS-CoV-2. Nat Rev Neurosci 2021;22(2):74–75. DOI: 10.1038/s41583-020-00427-3.

26. Yong SJ. Persistent Brainstem Dysfunction in Long-COVID: A Hypothesis. ACS Chem Neurosci 2021;12(4):573–580. DOI: 10.1021/acschemneuro.0c00793.

27. Song E, Zhang C, Israelow B, et al. Neuroinvasion of SARS-CoV-2 in human and mouse brain. J Exp Med 2021;218(3). DOI: 10.1084/jem.20202135.

28. Deleidi M, Isacson O. Viral and inflammatory triggers of neurodegenerative diseases. Sci Transl Med 2012;4(121):121ps3. DOI: 10.1126/scitranslmed.3003492.

