## Supplemental Figure 1 for "Undiagnosed COVID-19 in households with a child with mitochondrial disease"

**SUPPLEMENTAL DATA**

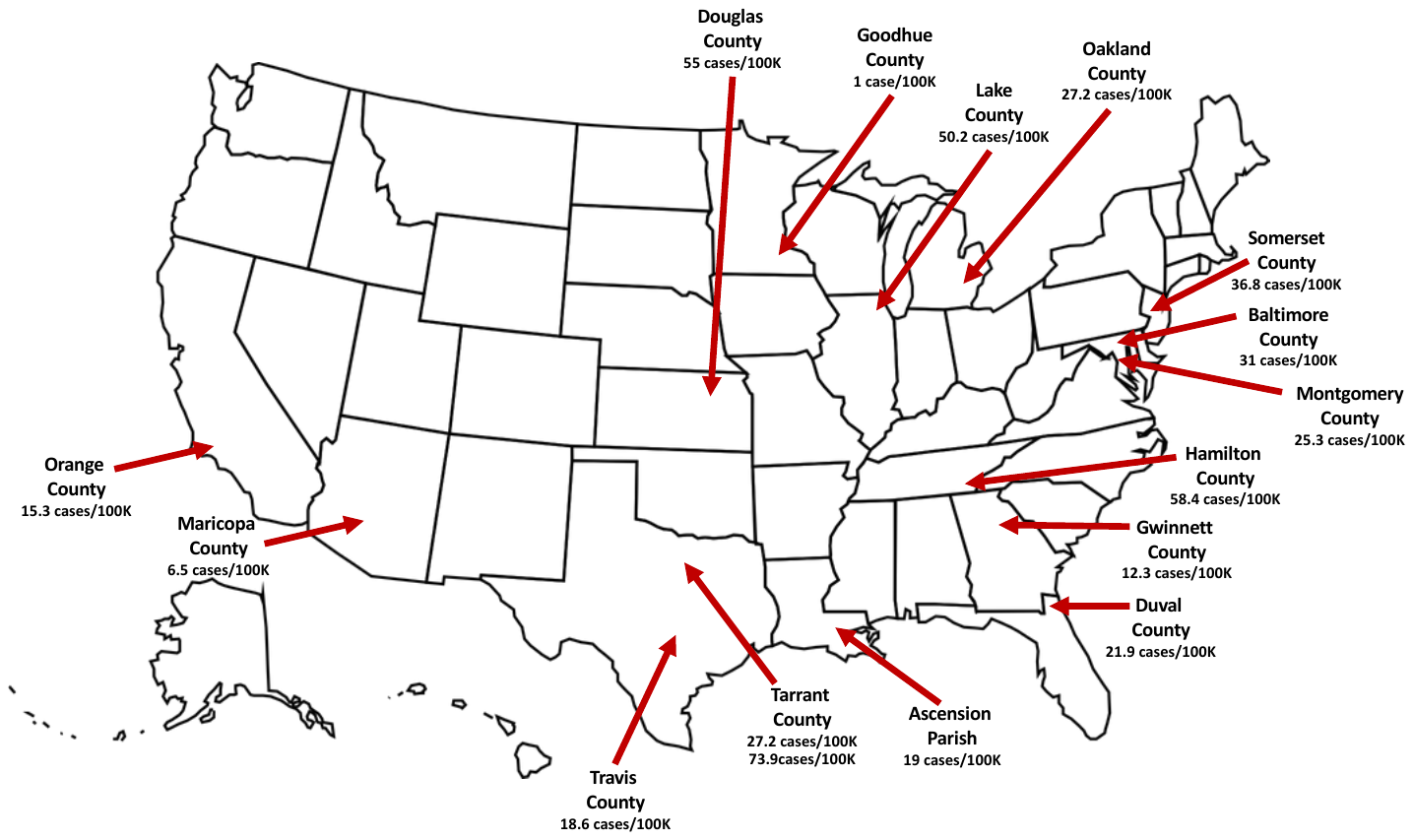

**Figure S1: Map of the United States showing number of cases by county.** Seven-day averages per 100K population were compiled for families on the day of collection of samples. Some counties (e.g., Tarrant County, TX) contained more than one family.
